# Timing of *S. aureus*-related mortality in a large randomized clinical trial: Implications for future study design

**DOI:** 10.64898/2026.06.20.26356148

**Authors:** Todd C. Lee, Guillaume Butler-Laporte, Matthew P. Cheng, Dominik Mertz, Ranjani Somayaji, Kevin Afra, Anthony Bai, Zain Chagla, Nick Daneman, Jennifer M. Grant, Jennie Johnstone, Christopher Kandel, Derek MacFadden, Sebastien Poulin, Connor Prosty, Kevin Schwartz, Michael Silverman, Stephanie Smith, Terence Wuerz, Steven Y.C. Tong, Emily G. McDonald

**Author notes:** Corresponding author: Todd C. Lee MD MPH, 1001 Decarie Blvd Room E5-1820, Montreal, QC, Canada, H4A3S1.

## Abstract

**Background:** Longer follow-up periods in clinical trials for *S. aureus* bacteremia (SAB) may capture unrelated deaths, adding random noise that risks biasing trial results towards the null.

**Objective:** To evaluate the timing and infection-relatedness of deaths within a large SAB clinical trial platform.

**Design:** Blinded duplicate adjudication of trial deaths using a modified 7-point Likert-Scale. A third reviewer settled disagreements.

**Setting:** 37 Canadian hospitals participating in the *S. aureus* Network Adaptive Platform (SNAP) Trial.

**Participants:** 1515 adult patients recruited to SNAP between February 2022 and May 2026.

**Measurements:** Timing and relatedness of 90-day deaths categorized as at least possibly SAB-related not likely to be SAB-related. Optimal follow-up cut-off was determined using Youden’s index and graphically.

**Results:** 247 deaths occurred; 97 (39.3%) were adjudicated as at least possibly SAB-related and 150 (60.7%) as not likely related. For probably/definitely related deaths, interrater agreement was 85.0% (Gwet’s AC=0.73, substantial); for at least possibly related, it was 77.3% (Gwet’s AC=0.55, moderate). Median survival was significantly shorter for SAB-related deaths (12 vs. 30.5 days; difference: 19 days earlier, 95% CI: 12-26, p<0.0001). Nearly 80% of SAB-related deaths occurred by day 30, whereas 50% of unrelated deaths occurred between days 30 and 90. Youden’s index optimized follow-up at 20.5 days.

**Limitations:** Potential for cause-of-death misclassification and data limited to Canadian sites.

**Conclusion:** Deaths considered attributable to SAB cluster rapidly within the first month, while later deaths are predominantly unrelated. A 30-day all-cause mortality window may be more appropriate than 90 days for primary mortality outcomes in trials evaluating acute SAB therapies with longer follow up reserved for metastatic infection and recurrence.

## Introduction

While mortality in *S. aureus* bacteremia has been decreasing over time (1), it remains a common cause of infectious death worldwide (2), with an associated all-cause mortality over 1 in 4 at 90-days. Randomized clinical trials (RCTs), such as the *S. aureus* Network Adaptive Platform (3) (SNAP), are working to standardize and optimize diagnosis and treatment approaches with the goal of improving outcomes in this lethal disease (4,5).

When designing an RCT, selection of the most appropriate primary outcome is critical. In 2017, a group of international experts proposed that a key primary outcome for phase 3 or 4 trials of *S. aureus* bacteremia could be defined as survival in the absence of microbiological failure at 90-days (6). As mortality may not capture other clinically relevant effects of different therapies, it was proposed that mortality be used alongside clinically meaningful secondary outcomes. Accordingly, the SNAP investigators chose all-cause mortality at 90-days as the primary outcome (3) with a variety of secondary outcomes encompassing efficacy and safety.

However, it has previously been suggested that longer follow-up times in *S. aureus* bacteremia, while increasing the numbers of overall deaths, may do so at the expense of including unrelated deaths (7). In that systematic review of 24 observational studies, the authors showed that 1 month of follow-up would capture the majority of *S. aureus* related mortality. In a large enough trial, or a trial where disease related deaths occur on a longer timeline, a small number of unrelated deaths is less likely to have a significant impact on the overall study conclusions. However, in a bloodstream infection trial where deaths due to acute infection generally occur over a very short time (days to weeks), the contribution of unrelated deaths occurring after the point where the infection has most likely been cured risks biasing the results towards the null. This could mean potentially either discarding a promising therapy or leading to a false conclusion of non-inferiority. To further explore this issue as it pertains to *S. aureus* bacteremia and to clinical trials, we adjudicated the timing and relatedness of deaths among Canadian SNAP trial participants.

## Methods

SNAP (clinicaltrials.gov NCT05137119) is an international platform trial which has been evaluating a variety of therapeutic options for *S. aureus* bacteremia since 2022 (3). This was a pre-planned analysis of adult participant data from the 37 Canadian SNAP sites. All participating sites submitted an anonymized clinical case summary of the circumstances of death for each participant from February 1, 2022, to May 11, 2026. This case summary was created by research personnel and site investigators by synthesizing the available clinical documentation into a summary paragraph. Details of treatment assignment and the elapsed time between recruitment and death were excluded from the reports.

### Evaluation of Timing and Relatedness of Death

Patients included in this analysis were adults who were recruited to the SNAP clinical trial platform within 72 hours of their index blood culture and who met the pragmatic inclusion criteria for at least one of SNAP’s domains (3). Using a web based platform, and building on our previous work in adverse drug events (8), the summary of each Canadian SNAP participant who died was reviewed independently, in duplicate, while blinded to timing of death and treatment assignment. Five reviewers, composed of 2 general internists and 3 infectious diseases specialists, were each randomly assigned a subset of the case files such that each file had two reviewers and the overlap between any two reviewers was at random. For each death, the reviewers rated the cause on a 7 point Likert scale based on the Leape and Bates adjudication method for Adverse Drug Events (9) (**Box 1**).

Cases received a final adjudication as probably or definitely related to *S. aureus* bacteremia if both of the reviewers gave the case file a score of 6 or 7 and as possibly related if both reviewers gave a score of 5, 6, or 7. Cases were adjudicated as not likely related to *S. aureus* bacteremia if both gave scores of 4 or below. In cases where there was categorical disagreement (one possible [>=5] and one not likely related [<=4]), a third reviewer, who was not one of the initial five, provided the tie-breaking opinion. Interrater agreement was evaluated by Gwet’s AC (10). The number of days between SNAP platform entry and death was used for the timing of death. For context, patients entered the SNAP platform at a median of 50-52 hours after the index blood culture and 6.3% of patients were excluded from the platform because they had already died when the index culture resulted (5).

### Analysis

All analyses were conducted using Stata version 17.0 (StataCorp, USA). First, we plotted the timing of probably/definitely related, possibly related, and unrelated deaths. The difference in median survival between these groups was calculated using quantile regression. We then visualized the cumulative incidence function for both related and unrelated deaths accounting for competing risks and using the 1515 Canadian patients recruited to SNAP as of May 1st as the denominator.

Next, used the Stata modules *cutpt* (11) and *roctab* to calculate the positive likelihood ratio for related vs. unrelated deaths as a function of follow-up time and the Youden index for the time point which found the time point which optimized the area under the receiver operating curve which best classified the data into these two groups. The Youden index and positive likelihood ratio were visualized as a function of follow-up time.

Finally, to provide an alternative visualization, we created a stacked bar graph based on various follow-up times and showing the cumulative proportion of deaths which were probably/definitely related, at least possibly related, and unrelated as a function of time.

## Results

As of May 1, 2026, there were a total of 1515 Canadian patients recruited to the SNAP platform. Overall, there were 247 deaths (16.3%) reported by day 90 among Canadian SNAP participants. Of these, 71 deaths (28.7%) were classified as probably/definitely *S. aureus* bacteremia related, 26 (10.5%) as at least possibly related, and 150 (60.7%) as not likely related. For assigning a death as being probably/definitely related to *S. aureus*, percent agreement between the initial evaluators was 85.0% with a Gwet’s AC 0.73 (Substantial agreement (12)) and for at least possibly related percent agreement was 77.3% with a Gwet’s AC of 0.55 (Moderate agreement). Fifty-six (22.7%) of cases required a third review. For these third reviews, 49 (87.5%) were subsequently scored as not likely related.

The median age of patients classified as at least possibly *S. aureus* related was 76 (IQR 66-84) and as not likely related was 75 (IQR 66-82) and 60% of patients were male with similar proportions. Median survival in those identified as at least possibly dying of *S. aureus* bacteremia was 12 days (IQR 4-20) versus 30.5 days (IQR 14-52) in those who were deemed not likely to be related. This corresponded to a difference of 19 days earlier (95%CI 12 to 26; p<0.0001). The cumulative incidence of death accounting for competing risk is shown in **Figure 1**. Because the curves of possibly related and probably/definitely related were virtually superimposable, these groups were combined. The cumulative incidence curves of at least possibly related and not likely related crossed at approximately day 30. By day 30, nearly 80% of all of the possibly related deaths had occurred compared to only 50% of not likely related deaths. Between day 30 and day 90, the remaining 20% of at least possibly related deaths occurred, whereas this time yielded an additional 50% of not likely related deaths.

**Figure 1.**
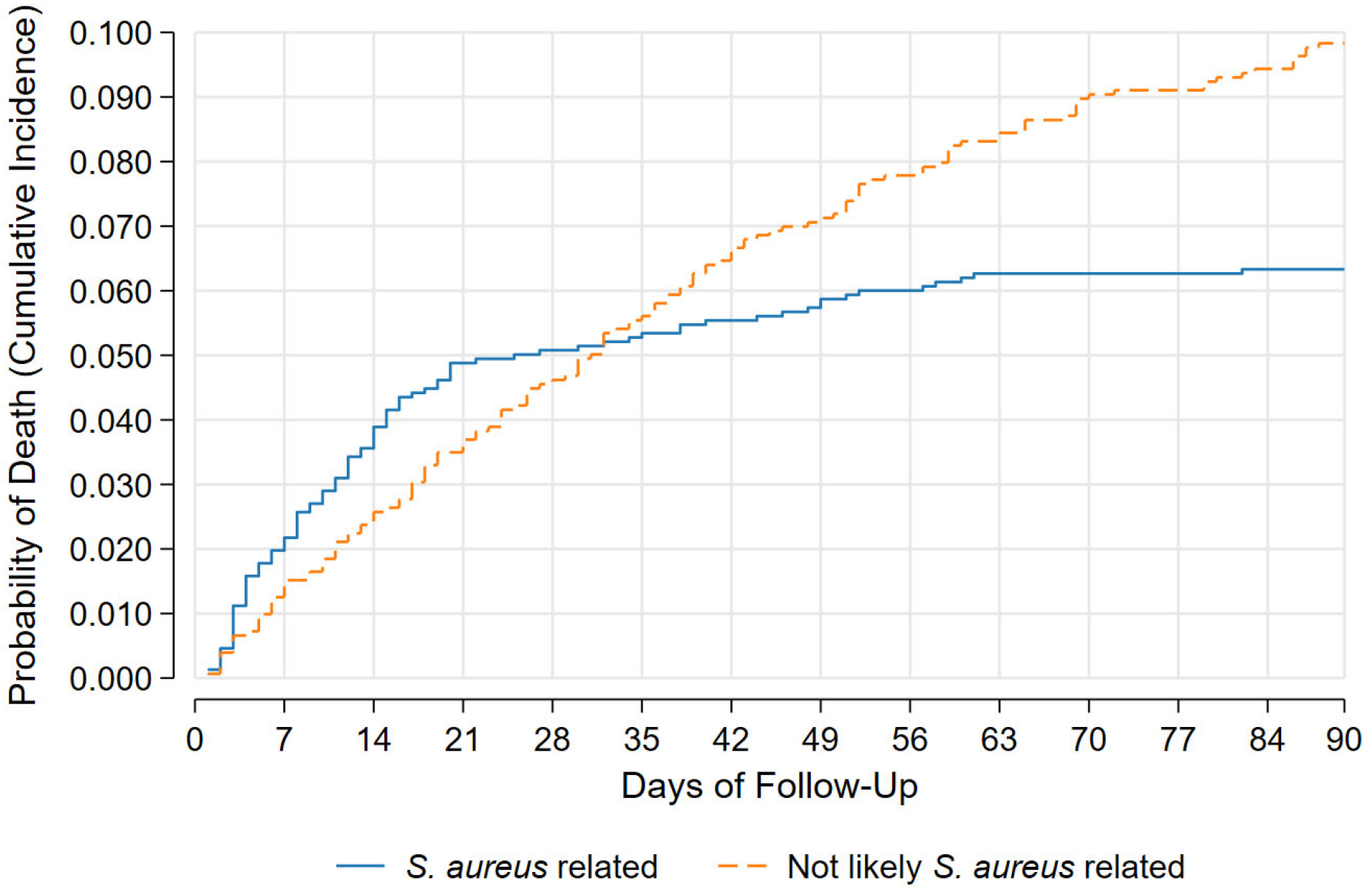
Cumulative Incidence of Mortality Accounting for Competing Risk

The optimal cut-off by Youden’s index was 20.5 days, with a sensitivity of 77% and specificity of 65%. The area under the curve was 0.71. The relationship between Youden’s index and time is shown in **Figure 2**. Extending follow-up to 30 days increased the sensitivity to 81.1%, decreased the specificity to 50%, but maintained a positive likelihood ratio of 1.6 that death was at least possibly related to *S. aureus* bacteremia. Beyond 30 days, the likelihood ratio approached 1, as incremental gains in sensitivity were exceeded by larger losses in specificity.

**Figure 2.**
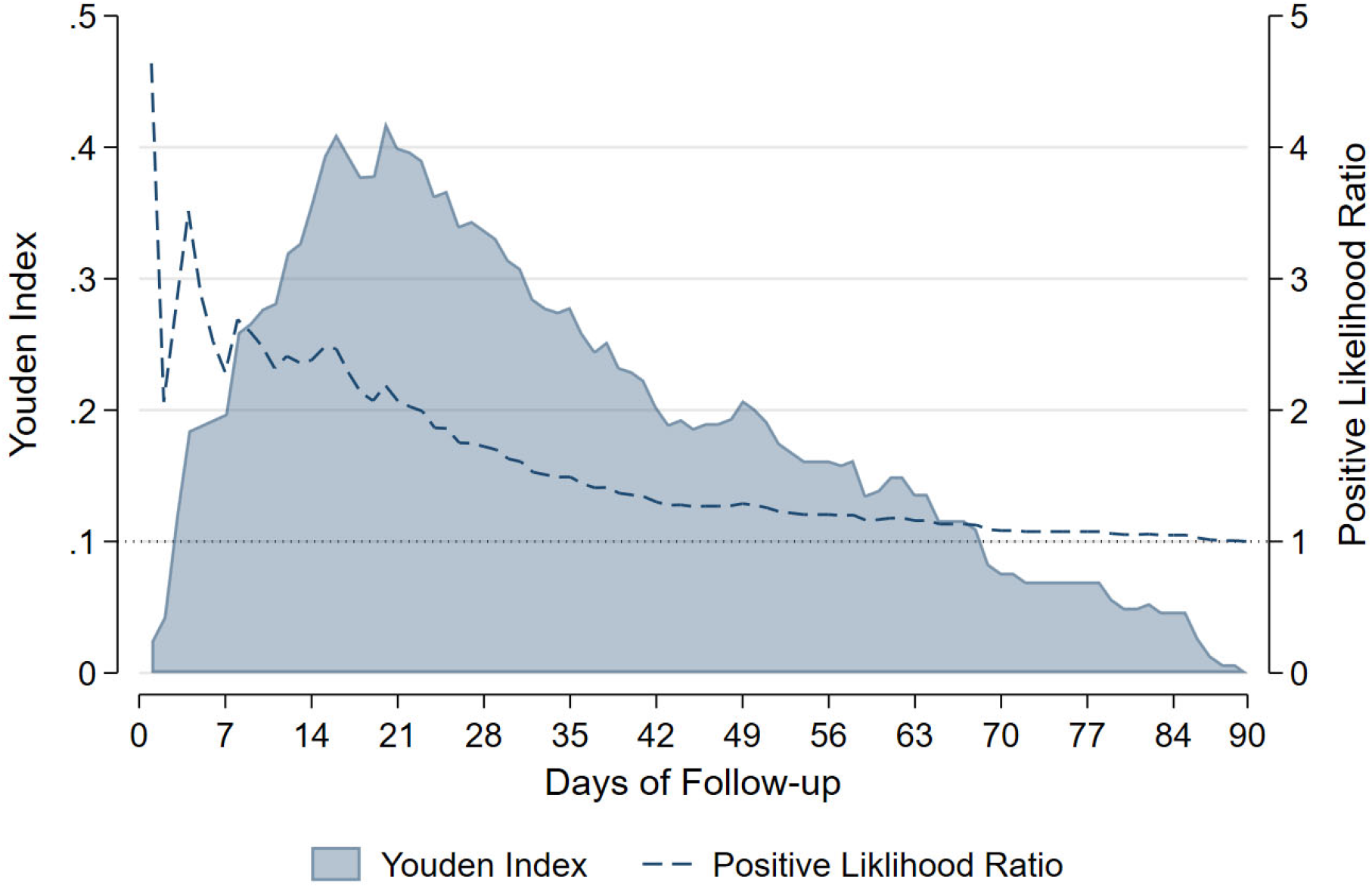
Youden index and positive likelihood ratio over time

The proportion of deaths which were probably/definitely related, possibly related, and not related as a function of time are shown in **Figure 3**. As follow-up time increased, the proportion of deaths felt not to be likely due to *S. aureus* increased until it reached 60%of all deaths by day 90.

**Figure 3.**
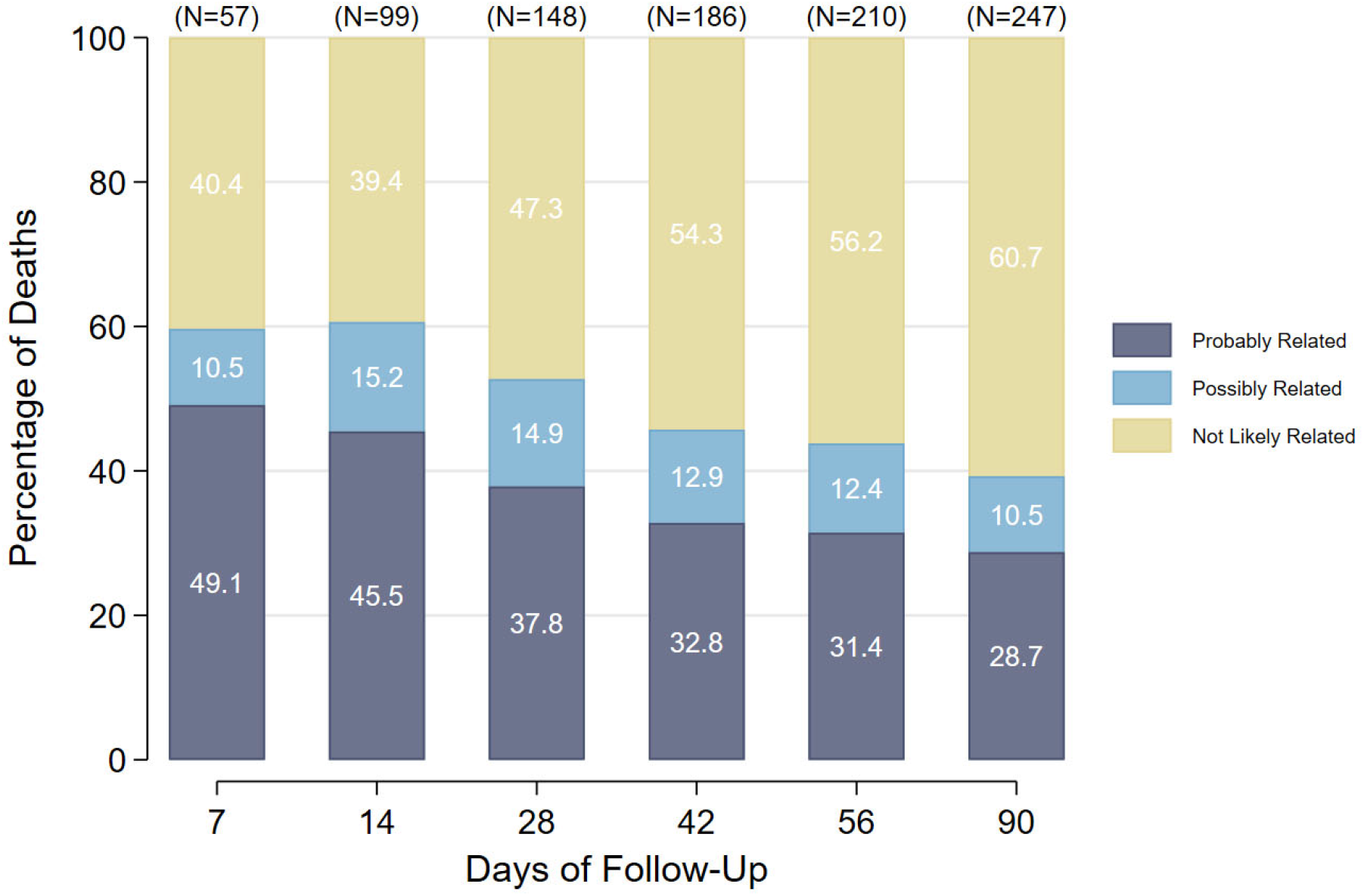
Proportion of deaths related to *S. aureus* over time.

## Discussion

In this preplanned analysis of deaths from the Canadian participants of the SNAP trial, we demonstrated that deaths that were adjudicated as being related to *S. aureus* bacteremia were more likely to occur earlier in follow-up. As follow-up time increased, so too did the number deaths which were not likely to be related. Based on our findings, bolstered by the complementary prior systematic review (7), it can be hypothesized that deaths due to the infection tend to occur rapidly and cluster earlier, whereas unrelated deaths demonstrate a more linear distribution throughout the extended follow-up period. This may have important implications for future randomized clinical trial designs in *S. aureus* bacteremia, and perhaps all bacteremia research. While a longer follow-up yields higher total event rates, and indeed more completely captures all related deaths, it also dilutes the specificity of the relatedness of the outcome and, paradoxically through the addition of random noise, could lead erroneous conclusions leading to discarding effective therapies or even adopting ineffective ones by chance. Indeed, in the previous systematic review (7), the authors simulated the effect of non-related deaths after 1 month in a hypothetical trial and showed an example of a trial which would be statistically significant at 30 days but non-significant at day 90.

Undoubtedly, all-cause mortality is of paramount importance as adjudication of cause of death is imperfect and adds complexity. Furthermore, a patient cannot die of *S. aureus* bacteremia if they die from treatment toxicity. However, if the goal is to identify therapeutics which reduce mortality in cases of *S. aureus* bacteremia, narrowing the follow-up window would more specifically capture related deaths without the challenges of adjudication. An important consideration is that death from *S. aureus* bacteremia (and indeed other infections) can occur through a variety of mechanisms: direct virulence of the bacteria; the host response; treatment toxicity; interactions between comorbidities, frailty, and the underlying infection; and hospital related iatrogenesis. Death over a longer period potentially captures all these contributors. Nonetheless, once a patient is cured, and the potential harm from drug-side effects has dissipated, deaths due to patient factors and competing risk may bias the study towards the null and reduce the chances to detect a potentially valid therapeutic option.

As medicine and care evolve, death from drug toxicities, host response, and iatrogenesis may decline through novel and alternative strategies. Based on the data we present, we believe that 30-day follow-up is likely a more ideal period for the primary outcome of all-cause death when comparing regimens for the acute therapy of *S. aureus* bacteremia, with longer follow-up for secondary outcomes including microbiological or treatment failure and late drug toxicities. Conversely, for questions of total duration of therapy or oral switch (13), where patients have (a) clinically improved and (b) already survived 2-4 weeks (the period where ~80% of *S. aureus* related death would have occurred), longer follow-up with a composite outcome which includes mortality and microbiologic failure may become the prudent choice.

Two examples from trials where our findings have important implications are worth discussing. For example, in the ARREST trial (14), which compared adjunctive rifampin to placebo in *S. aureus* bacteremia, early mortality in the rifampin group was higher in the first 28-36 days, with the survival curves converging towards 84 days. A harm signal from rifampin was potentially diluted by the longer follow-up. This is particularly relevant when the duration of rifampin for most patients was 2 weeks and afterwards any harms from the drug would be expected to dissipate as it was metabolized. Another example comes from the international SNAP trial (5), where the comparison of cefazolin to (flu)cloxacillin was 99.2% probable to be superior for all-cause mortality at day 28, but reduced to 89% by the day 90 primary outcome. Here, it is possible that later unrelated deaths may have prevented the trial from demonstrating the superiority of cefazolin.

### Limitations

This study has several important limitations. Most notably, any attribution of death in clinical medicine and in research has the potential for misclassification and this misclassification may differ based on how long after a potential inciting event death may have occurred. There may also be challenges in cases where there is potential for multicausality. Nonetheless, as the current 30- and 90-day follow-up times are based on expert opinion, we believe our data provides important empirical evidence for the discussion. Secondly, there was no established and universally accepted standard method to evaluate causality of death for *S. aureus* bacteremia and therefore we chose to modify an approach from the adverse drug event literature using a Likert-scale approach. Nonetheless, we believe this approach is valid given the moderate and substantial agreement in first reviews, the use of the third reviewer to blindly settle disagreements, and the fact that the method is transparent and reproducible. Another limitation is that we only had data from the Canadian sites from the SNAP trial. While our findings may be less generalizable versus the international SNAP study, our estimate for 30 day attributable mortality was similar to those found in the prior systematic review of observational studies (7). Finally, SNAP could not include the sickest patients who died prior to their blood culture result being available and so we likely underestimate early attributable deaths when compared to observational studies (15).

## Conclusion

In this analysis of Canadian participants of the SNAP trial, we show that deaths that were considered likely to be *S. aureus* related occur earlier than those which were considered not likely. We believe these findings have implications for future clinical trial design in *S. aureus* bacteremia. Our data suggest that 30-day mortality may be a more appropriate timepoint for future RCTs which evaluate aspects of the acute management of *S. aureus* bacteremia, with longer follow-up being used for secondary endpoints. In circumstances where most of the disease attributable mortality has already occurred at recruitment, it would likewise be prudent to use outcomes such as microbiological failure at longer time points.

## Data Availability

All data produced in the present work are contained in the manuscript.

## Acknowledgments

<u>Funding Source</u>: Funders had no role in the conduct or analysis of this RCT. The SNAP trial received funding from the Canadian Institutes of Health Research (#444369).

<u>IRB Approval</u>: All Canadian centres received institutional research ethics board approval for the conduct of SNAP and participants consented to secondary use of their data.

---

**Box 1:**
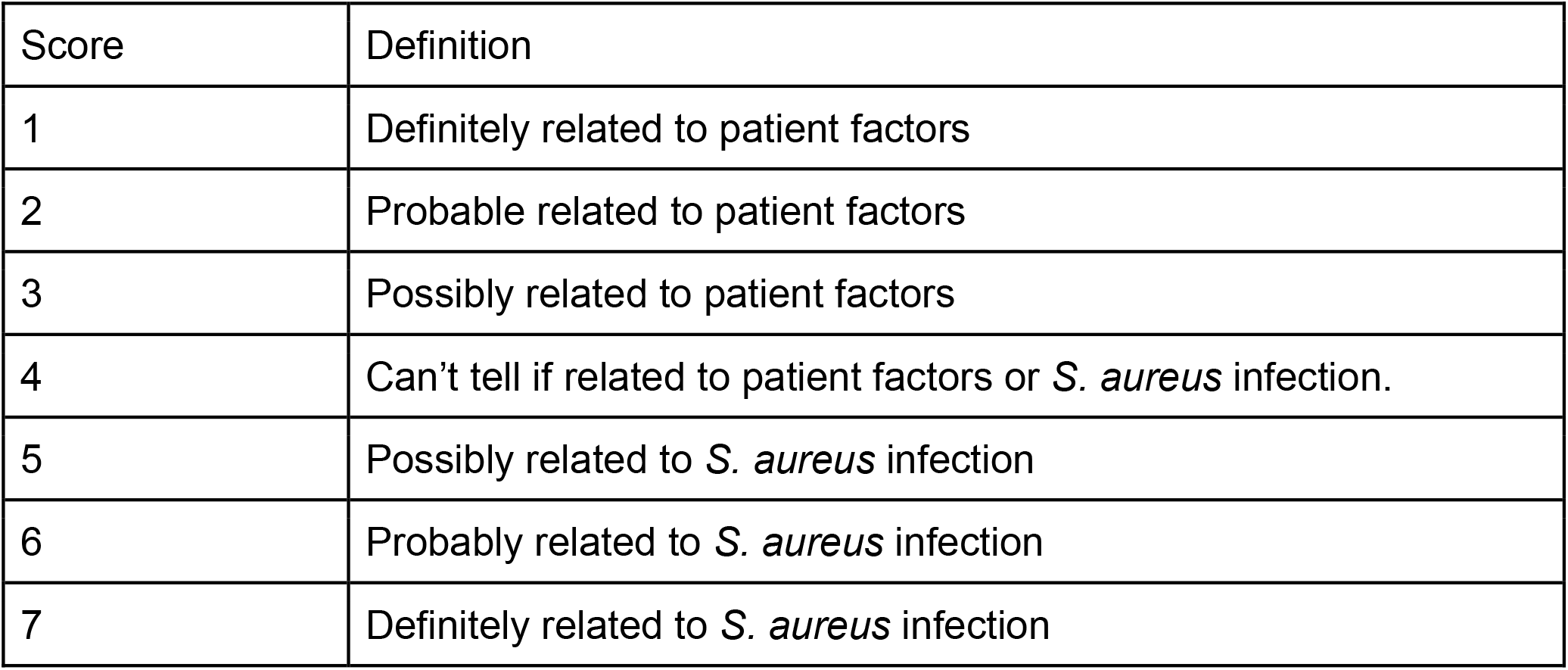
7-point Likert Scale to Evaluate Cause of Death.

**Box 1: 7-point Likert Scale to Evaluate Cause of Death**
| Score | Definition |
| --- | --- |
| 1 | Definitely related to patient factors |
| 2 | Probable related to patient factors |
| 3 | Possibly related to patient factors |
| 4 | Can't tell if related to patient factors or *S. aureus* infection. |
| 5 | Possibly related to *S. aureus* infection |
| 6 | Probably related to *S. aureus* infection |
| 7 | Definitely related to *S. aureus* infection |

